# Mitochondrial bioenergetic dysfunction and cryptic splicing of stathmin-2 are neuropathological markers of disease duration in sporadic amyotrophic lateral sclerosis

**DOI:** 10.1101/2022.09.01.22279305

**Authors:** Arpan R. Mehta, Karina McDade, Judith Newton, Marc-David Ruepp, Suvankar Pal, Siddharthan Chandran, Colin Smith, Bhuvaneish T. Selvaraj

## Abstract

A striking feature of sporadic amyotrophic lateral sclerosis (ALS) is the marked heterogeneity in disease duration; despite the stark median survival of three years from symptom onset, 10-20% of people with ALS survive longer than 10 years. An improved understanding of the mechanisms underpinning this is vital to revealing the biological basis of disease resilience. Accumulating experimental and pathological evidence implicates mitochondrial bioenergetic dysfunction and TDP-43 nuclear loss-of-function in the aetiopathogenesis of ALS. However, the relevance of these two molecular dysfunctions to disease duration and resilience in ALS is unknown. We curated a cohort of sporadic ALS cases comprising clinically linked autopsy samples to identify molecular neuropathological correlates of disease duration. We developed a novel dual BaseScope RNA in situ hybridisation probe that labels mitochondrial complex 1 transcript (MT-ND2) and truncated stathmin-2 (STMN2) transcripts to measure mitochondrial bioenergetic function and TDP-43 loss-of-function, respectively, in ventral horn neurons. We first show that there is dysfunctional mitochondrial bioenergetics in sporadic ALS. We observed reduced expression of MT-ND2 and increased expression of truncated STMN2 in ALS cases (N=20) compared to sex- and age-matched controls (N=10). We show that these findings correlate with ALS disease duration. Further mechanistic studies are needed to explore whether manipulation of STMN2 expression, by either suppressing cryptic splicing or overexpression, could modify disease duration.

---

A striking, but underappreciated, feature of amyotrophic lateral sclerosis (ALS) is the marked heterogeneity in disease duration (Brown and Al-Chalabi, 2017); despite the stark median survival of three years from symptom onset, 10–20% of people with ALS survive longer than 10 years (Chio *et al*., 2009). Improved understanding of the mechanisms underpinning these differences—in short and long survivors of ALS—is vital to inform both on the biological basis of disease resilience, as well as having implications for clinical trial design (Wong *et al*., 2021). Currently, there are no well-established molecular neuropathological correlates of disease duration.

Accumulating experimental and pathological evidence implicates an important role for mitochondrial dysfunction in the aetiopathogenesis of ALS (Dupuis *et al*., 2011; Dafinca *et al*., 2016; Vandoorne *et al*., 2018; Mehta *et al*., 2019; Smith *et al*., 2019; Allen, 2020; Dafinca *et al*., 2020; Sassani *et al*., 2020). We recently described a spinal motor neuron selective reduction in expression of mitochondrial electron transport chain transcripts in the *C9orf72* repeat expansion mutation—the commonest known genetic cause of ALS—as a driver of axonal dysfunction (Mehta *et al*., 2021). This leaves unanswered the wider relevance of this finding to sporadic ALS (sALS), which accounts for over 90% of all ALS cases, and its impact on disease duration. Early post-mortem studies of sporadic cases have yielded conflicting results, with evidence of reduced electron chain function in spinal cord (Fujita *et al*., 1996; Borthwick *et al*., 1999; Wiedemann *et al*., 2002; Ladd *et al*., 2017), whereas others have shown changes only in the motor cortex of *SOD1* cases and not in sALS (Bowling *et al*., 1993; Browne *et al*., 1998). Furthermore, a recent magnetic resonance spectroscopy study demonstrated bioenergetic dysfunction in sALS subjects (and an over-representation of bulbar disease), with evidence of defective mitochondrial oxidative phosphorylation in the brainstem (Sassani *et al*., 2020).

All sALS cases display the neuropathological hallmark of TDP-43 mis-localisation and accumulation, with both pathological (cytoplasmic) toxic gain-of-function and loss of normal (nuclear) function playing pathophysiological roles (Lee *et al*., 2011; Klim *et al*., 2019). Recent studies have shown that TDP-43 loss of function leads to downregulation of stathmin-2 (*STMN2*) mRNA in human motor neurons. This is due to the cryptic splicing and inclusion of a region within intron 1 (exon 2a) of *STMN2*, causing the introduction of a premature STOP codon and polyadenylation site. Consequentially, this leads to the expression of a truncated variant of *STMN2* mRNA that lacks exon 2-exon 5 (Klim *et al*., 2019; Melamed *et al*., 2019). Expression of this truncated *STMN2* mRNA variant is associated with TDP-43 cytoplasmic aggregation in frontotemporal dementia (Prudencio *et al*., 2020), supporting the case that TDP-43 pathology leads to nuclear loss-of-function of TDP-43. Although the extent of pathological TDP-43 burden does not correlate with disease duration (Brettschneider *et al*., 2013) or severity of non-motor phenotypes (Prudlo *et al*., 2016; Gregory *et al*., 2020), there is growing interest into the impact of TDP-43 proteinopathy on mitochondrial and axonal dysfunction in ALS (Wang *et al*., 2016; Izumikawa *et al*., 2017; Briese *et al*., 2020; Yu *et al*., 2020; Fazal *et al*., 2021; Zuo *et al*., 2021). Whether this relates to gain-of-function or loss-of-function remains unclear.

Against this background, and noting also recent findings of transcriptional dysregulation in pathways involved in mitochondrial function and energy production in TDP-43-depleted motor neurons (Briese *et al*., 2020), we set out to profile mitochondrial bioenergetic transcriptomic dysfunction and TDP-43 loss-of-function within the same motor neuron in ventral spinal cord human post-mortem tissue. Specifically, we wished to examine whether TDP-43 loss-of-function and mitochondrial bioenergetic function were associated with disease duration. We thus deployed a novel duplex BaseScope™ RNA *in situ* hybridisation (ISH) assay and designed probes against: (1) *MT-ND2* mRNA (as per Mehta *et al*. (2021)); and, (2) exon 2a of *STMN2* (probe name: BA-Hs-STMN2-E1E2A, Advanced Cell Diagnostics™ Bio-Techne) that selectively labels the truncated form of *STMN2* mRNA, as surrogates of mitochondrial electron transport chain (complex I) function and TDP-43 loss-of-function, respectively (Klim *et al*., 2019; Melamed *et al*., 2019; Mehta *et al*., 2021). We first confirmed specificity of the truncated *STMN2* probe by performing BaseScope™ ISH on induced pluripotent stem-cell derived spinal human motor neurons (Selvaraj *et al*., 2018; Mehta *et al*., 2021; Mehta *et al*., 2022), observing signal positivity in only TDP-43-depleted human motor neurons (generated according to (Reber *et al*., 2018; Mehta *et al*., 2022), **Supplementary Figure 1**). We used the Edinburgh Brain Bank cohort of sALS cases (curated from the Scottish national CARE-MND register), with no known ALS genetic mutations, of varying disease duration from symptom onset (*N* = 20; median disease duration = 46 months, interquartile range = 82 months; range of ages at death = 40 – 85 years, interquartile range = 12 years; ALS gene panel and *C9orf72* repeat-primed PCR negative as per Leighton *et al*. (2019) compared with age- and sex-matched controls (*N* = 10; range of ages at death = 57 – 75 years, interquartile range = 11 years) from the Sudden Death Brain Bank, with no neurological disorder during life and no significant neuropathology present at post-mortem (**Table 1**).

**Table 1:** Clinical meta-data for sporadic ALS cases used in post-mortem work. Cases 1 to 10 were classified as ‘short survivors’, whereas cases 11 to 20 were classified as ‘long survivors’, as per Westeneng *et al*. (2018). N.B. “Age at death” has been shown as a ‘range’ for anonymisation purposes, as per medRxiv policy. Abbreviations: UL – Upper limb; LL – Lower limb; F – Female; M – Male.

| Case ID<br>(Sex) | Age at<br>death<br>(years) | Disease<br>duration<br>(months) | Clinical<br>region<br>onset |
| --- | --- | --- | --- |
| Case 1<br>(F) | 65-70 | 5.60 | Cognition |
| Case 2<br>(M) | 55-60 | 11.40 | Bulbar |
| Case 3<br>(F) | 60-65 | 13.63 | UL & LL |
| Case 4<br>(M) | 55-60 | 14.63 | Bulbar |
| Case 5<br>(M) | 55-60 | 16.97 | Bulbar |
| Case 6<br>(M) | 65-70 | 17.37 | UL |
| Case 7<br>(M) | 60-65 | 18.60 | Bulbar |
| Case 8<br>(F) | 75-80 | 21.33 | Bulbar |
| Case 9<br>(M) | 75-80 | 22.73 | Bulbar |
| Case 10<br>(M) | 70-75 | 24.37 | Bulbar |
| Case 11<br>(F) | 70-75 | 68.40 | Bulbar |
| Case 12<br>(M) | 40-45 | 69.03 | Bulbar |
| Case 13<br>(M) | 55-60 | 71.00 | UL |
| Case 14<br>(M) | 65-70 | 93.80 | UL |
| Case 15<br>(F) | 40-45 | 98.90 | LL |
| Case 16<br>(F) | 80-85 | 100.07 | LL |
| Case 17<br>(M) | 70-75 | 103.87 | UL |
| Case 18<br>(M) | 60-65 | 113.33 | UL |
| Case 19<br>(F) | 85-90 | 165.33 | LL |
| Case 20<br>(M) | 70-75 | 219.67 | UL |

We observed significantly reduced mRNA expression of *MT-ND2* and increased expression of truncated *STMN2* in ventral horn spinal motor neurons in sALS (**Fig 1**). The expression of truncated *STMN2* mRNA was specific to sALS, as opposed to other neurodegenerative diseases, such as Alzheimer’s dementia and Parkinson’s disease with Lewy body dementia (**Supplementary Fig 2**). We next examined whether these findings had a relationship with disease duration using our sALS cohort, stratifying cases into those with a short disease duration from symptom onset to death (*N* = 10; median disease duration = 17 months, interquartile range = 7 months; range of ages at death = 57 – 77 years, interquartile range = 12 years) *versus* long disease duration (*N* = 10; median disease duration = 99 months, interquartile range = 34 months; range of ages at death = 40 – 85 years, interquartile range = 14 years), as per Westeneng *et al*. (2018). We observed that the short survivors had significantly lower expression of *MT-ND2* and higher expression of cryptic *STMN2* than the long survivors (**Fig 2a,b**). Indeed, the positive and negative correlations between *MT-ND2* expression and disease duration, and truncated *STMN2* expression and disease duration, respectively, are significant (**Fig 2c**), suggesting a strong biological gradient and association.

**Fig 1:**
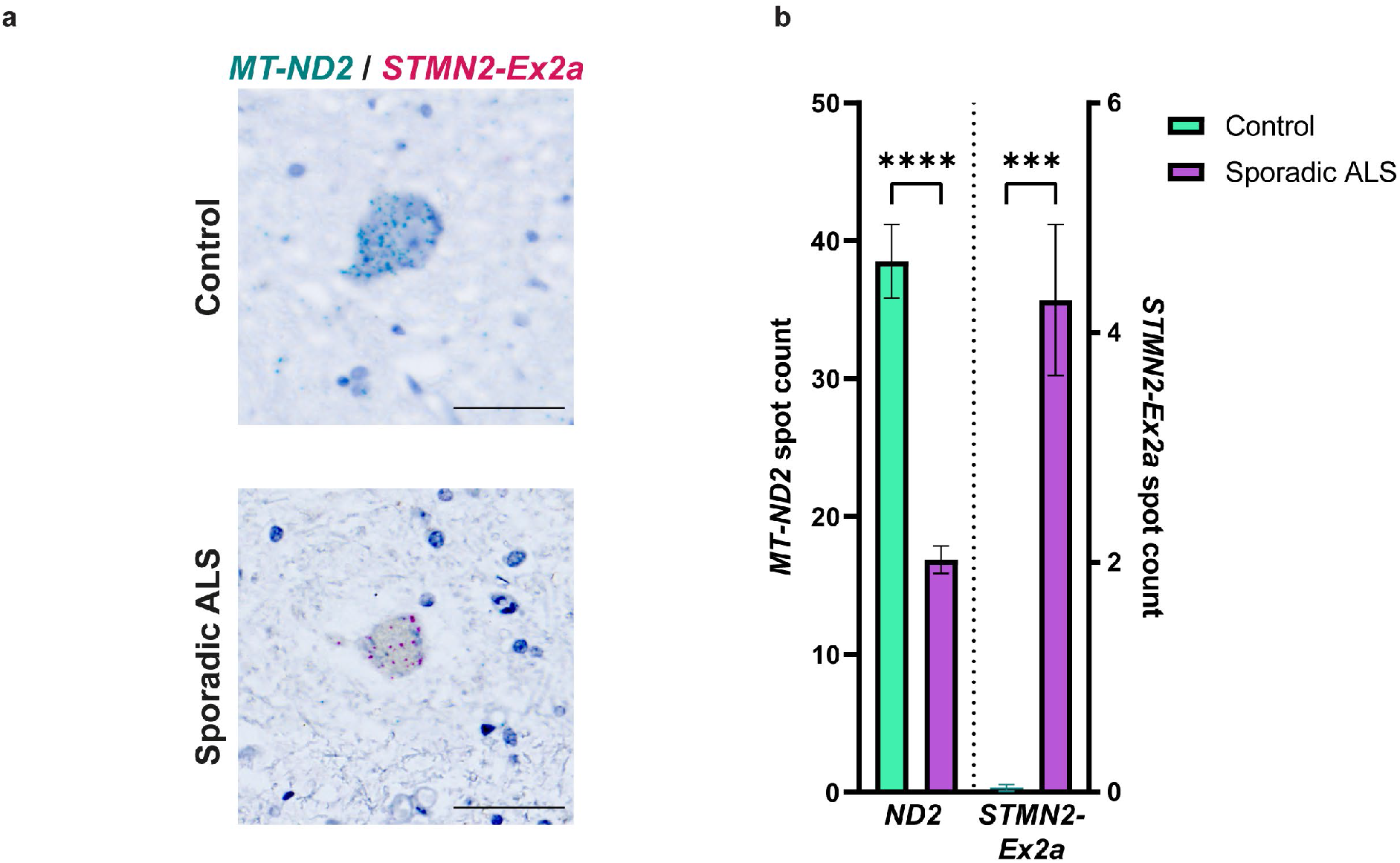
Sporadic ALS displays transcriptional markers of mitochondrial bioenergetic dysfunction and TDP-43 loss of function. **a**, Representative photomicrographs of ventral horn spinal motor neurons in healthy control *versus* sporadic ALS case, examined using a BaseScope™ dual probe that recognises individual mRNA molecules of *MT-ND2* (teal coloured spots) and truncated *STMN2* (‘*STMN2-Ex2a’*; red coloured spots). Tissue was counterstained with haematoxylin. Scale bars = 50 µm. **b**, Bar chart depicting the quantification of the number of *MT-ND2* (left ordinate axis) and truncated *STMN2* (right ordinate axis) transcripts per ventral horn neuron. Bars represent aggregate mean ± S.E.M. for sporadic ALS (purple bars; *N* = 20 cases; *n* = 2-12 evaluated motor neurons per case) and their age- and sex-matched controls (green bars; *N* = 10; *n* = 2-12). Statistical significance was evaluated with the Kruskal-Wallis test with a 95% Benjamini-Hochberg false discovery rate correction for multiple comparisons. ^*^*p* < 0.05, ^**^*p* < 0.01, ^***^*p* < 0.001, ^****^*p* < 0.0001.

**Fig 2:**
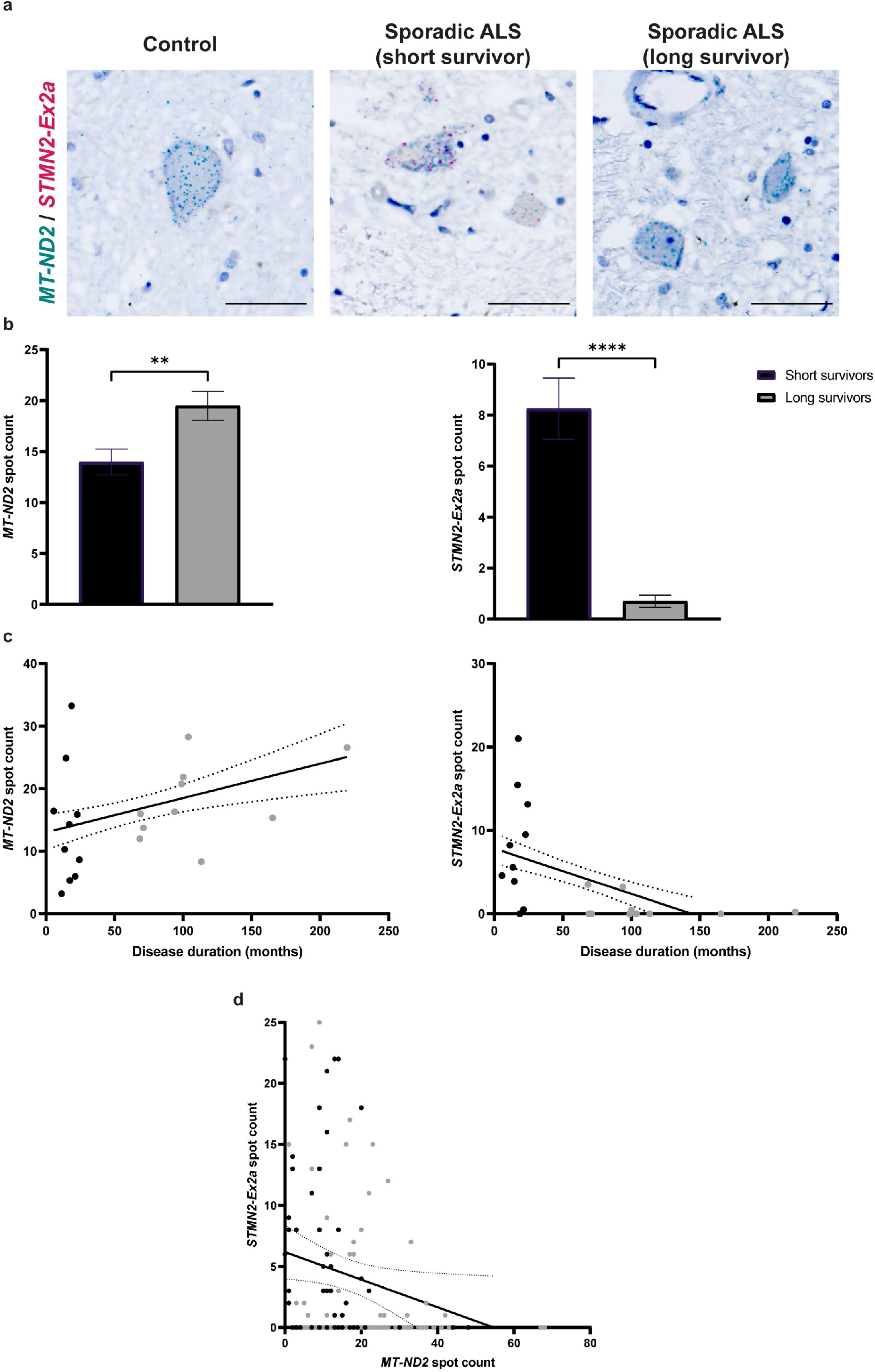
The extent of motor neuronal mitochondrial bioenergetic dysfunction and loss of TDP-43 function distinguishes long survivors of sporadic ALS from short survivors. **a**, Representative photomicrographs of ventral horn spinal motor neurons in healthy control *versus* short survivor sporadic ALS case (disease duration: 17 months), *versus* long survivor sporadic ALS case (disease duration: 104 months) examined using a BaseScope™ dual probe that recognises individual mRNA molecules of *MT-ND2* (teal coloured spots) and truncated *STMN2* (‘*STMN2-Ex2a’*; red coloured spots). Tissue was counterstained with haematoxylin. Scale bars = 50 µm. **b**, Bar chart depicting the quantification of the number of *MT-ND2* (left) and truncated *STMN2* (right) transcripts per ventral horn neuron. Bars represent aggregate mean ± S.E.M. for short survivor cases of sporadic ALS (black bars; *N* = 10 cases; *n* = 2-12 evaluated motor neurons per case) *versus* long survivor cases of sporadic ALS (grey bars; *N* = 10 cases; *n* = 2-12 evaluated motor neurons per case). Statistical significance was evaluated with the Kruskal-Wallis test. ^*^*p* < 0.05, ^**^*p* < 0.01, ^***^*p* < 0.001, ^****^*p* < 0.0001. **c**, Scatterplots assessing the correlation between disease duration (abscissa) *versus MT-ND2* spot count (ordinate; left) or truncated *STMN2* spot count (ordinate; right) in ventral horn spinal motor neurons. Data points in short sporadic ALS survivors (*N* = 10 cases; *n* = 2-12 evaluated motor neurons per case) and long survivors (*N* = 10 cases; *n* = 2-12 evaluated motor neurons per case) are depicted in black and grey colour, respectively. Simple linear regression best fit line and 95% confidence bands are shown (*p* = 0.0011 for *MT-ND2* spot count/disease duration correlation; *p* < 0.0001 for truncated *STMN2* spot count/disease duration correlation). **d**, Scatterplots assessing the correlation between *MT-ND2* spot count (abscissa) *versus* truncated *STMN2* spot count (ordinate) in ventral horn spinal motor neurons. Data points for short sporadic ALS survivors (*N* = 10 cases; *n* = 2-12 evaluated motor neurons per case) and long survivors (*N* = 10 cases; *n* = 2-12 evaluated motor neurons per case) are depicted in black and grey colour, respectively. Simple linear regression best fit line (*p* = 0.0361) and 95% confidence bands are shown.

Lastly, we exploited the duplex BaseScope™ ISH assay that facilitates the simultaneous assessment of *MT-ND2* and truncated *STMN2* mRNA in the same ventral horn neurons to assess if there is an association between TDP-43 loss-of-function and mitochondrial bioenergetic dysfunction. We observed that the expression of truncated *STMN2* mRNA was negatively associated with the expression of *MT-ND2* (**Fig 2d**), suggesting that TDP-43 loss- of-function might play a role in transcriptional regulation of mitochondrial bioenergetics.

In summary, we show in human post-mortem tissue that perturbed mitochondrial bioenergetics is a prominent feature of sALS, correlating with disease duration. Whilst truncated *STMN2* positivity has recently been shown in human post-mortem tissue to be associated with an earlier age of onset in frontotemporal dementia (Prudencio *et al*., 2020), there had hitherto been no relationship with disease duration demonstrated in sALS. This is further corroborated by recent findings that genetic polymorphism in *STMN2* is associated with bulbar onset and shorter disease duration (Theunissen *et al*., 2021). Our data provide a rationale for the rapid development of liquid biopsy RNA or protein biomarkers focussed on mitochondrial dysfunction and truncated *STMN2*. In addition, further mechanistic studies will be important in determining whether manipulation of *STMN2* expression, by either suppressing cryptic splicing or overexpression, could modify disease duration.

## Data Availability

All data produced in the present study are available upon reasonable request to the authors.

## Acknowledgements

The authors thank: (i) Edinburgh Brain Bank for supplying all post-mortem brain material and the Scottish MND Register/CARE-MND Consortium for all clinical and demographic data; (ii) Scottish MND clinical specialist team for discussing and obtaining consent from patients with ALS/MND for inclusion in these resources; and, (iii) MND Scotland and the Sylvia Aitken Charitable Trust for funding C.S. to help establish the MND Tissue Bank. Ethical approval was granted by the Scotland A Research Ethics Committee for the Scottish Clinical Audit Research Evaluation for Motor Neuron Disease platform (CARE-MND; 15/SS/0216), now a sub-registry of Rowling CARE (16/SS/0156; IRAS ID 200777), for this and related studies.

## Author contributions

A.R.M., S.C., C.S. and B.T.S. conceived and designed the study. K. McD. performed the post-mortem tissue work from a cohort curated by J.N. and S.P. M.D.R contributed to the methodology and resources. C.S. performed blinded neuropathological data collection. A.R.M. and B.T.S. performed data analysis. C.S. and S.C. provided resources and contributed to the interpretation of data with A.R.M. and B.T.S. A.R.M. drafted the manuscript, with critical input from S.C., C.S. and B.T.S. All authors revised and approved the final manuscript.

