## Supplementary Material for "Mitochondrial bioenergetic dysfunction and cryptic splicing of stathmin-2 are neuropathological markers of disease duration in sporadic amyotrophic lateral sclerosis"

**Supplementary Figures 1-2**

**Supplementary Table 1**

**Supplementary Fig 1**

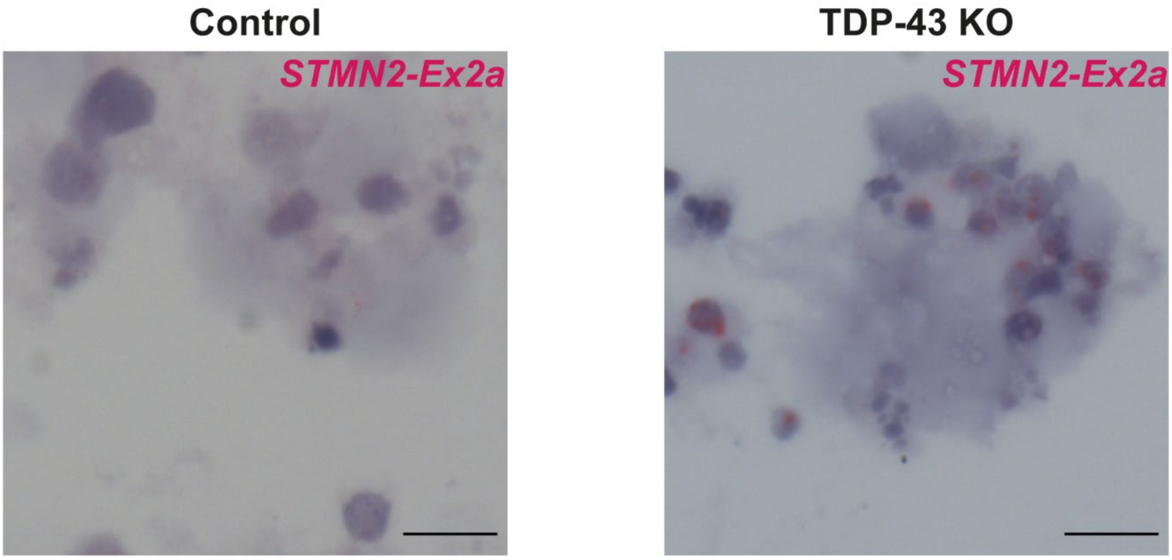


**Supplementary Fig 1: Specificity of BaseScope™ RNA *in situ* hybridisation probe recognising truncated *STMN2* mRNA.** Representative photomicrographs of an induced pluripotent stem-cell derived control spinal motor neuron (left) *versus* TDP-43 depleted spinal motor neuron (right). Each red coloured spot denotes an individual mRNA molecule of truncated *STMN2* (‘*STMN2-Ex2a’*), seen only in TDP-43 depleted cells. Motor neurons were counterstained with haematoxylin. Scale bars = 10 μm.

**Supplementary Fig 2**

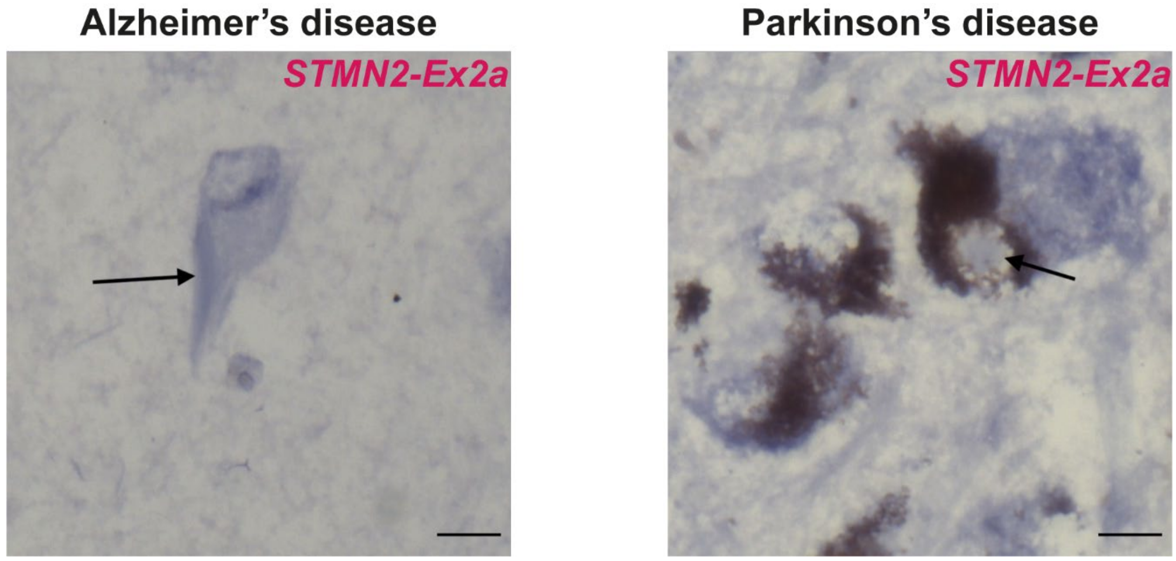


**Supplementary Fig 2: The expression of truncated *STMN2* mRNA is specific to TDP-43 pathophysiology.** Representative photomicrographs of a neuron from a post-mortem case of Alzheimer's disease (left) *versus* Parkinson's disease (right), neither of which harbour detectable neuronal truncated *STMN2* mRNA ('*STMN2-Ex2a*') via the BaseScope™ *in situ* hybridisation assay (absence of red coloured spots in both cases). The arrow refers to a neurofibrillary tangle in the case of Alzheimer's disease, and a Lewy body in the case of Parkinson's disease. Neurons were counterstained with haematoxylin. Scale bars = 10 µm.

**Supplementary Table 1: Age- and sex-matched controls.**

N.B. “Age at death” has been shown as a ‘range’ for anonymisation purposes, as per medRxiv policy.

Abbreviations: F – Female; M – Male.

| Control ID (Sex) | Age at death (years) |
|---|---|
| 1 (M) | 55-60 |
| 2 (M) | 55-60 |
| 3 (F) | 55-60 |
| 4 (F) | 60-65 |
| 5 (M) | 60-65 |
| 6 (F) | 65-70 |
| 7 (M) | 65-70 |
| 8 (M) | 70-75 |
| 9 (F) | 70-75 |
| 10 (M) | 75-80 |
